# Sex and gender differences in COVID testing, hospital admission, presentation, and drivers of severe outcomes in the DC/Maryland region

**DOI:** 10.1101/2021.04.05.21253827

**Authors:** Eileen P. Scully, Grant Schumock, Martina Fu, Guido Massaccesi, John Muschelli, Joshua Betz, Eili Y. Klein, Natalie E. West, Matthew Robinson, Brian T. Garibaldi, Karen Bandeen-Roche, Scott Zeger, Sabra L. Klein, Amita Gupta, for the JH-CROWN registry team

**Affiliations:** Department of Medicine, Division of Infectious Diseases, Johns Hopkins University School of Medicine, Baltimore, Maryland, USA; Department of Biostatistics, The Johns Hopkins Bloomberg School of Public Health, Baltimore, Maryland USA; Department of Emergency Medicine, Johns Hopkins University School of Medicine; Division of Pulmonary and Critical Care Medicine, Johns Hopkins University School of Medicine; W. Harry Feinstone Department of Molecular Microbiology and Immunology, The Johns Hopkins Bloomberg School of Public Health, Baltimore, Maryland USA; Department of Biochemistry and Molecular Biology, The Johns Hopkins Bloomberg School of Public Health, Baltimore, Maryland USA; Department of International Health, The Johns Hopkins Bloomberg School of Public Health, Baltimore, Maryland USA

**Keywords:** BMI, CRP, kidney disease, pulmonary disease, tocilizumab

## Abstract

**Background:** Rates of severe illness and mortality from SARS-CoV-2 are greater for males, but the mechanisms for this difference are unclear. Understanding the differences in outcomes between males and females across the age spectrum will guide both public health and biomedical interventions.

**Methods:** Retrospective cohort analysis of SARS-CoV-2 testing and admission data in a health system. Patient-level data were assessed with descriptive statistics and logistic regression modeling was used to identify features associated with increased male risk of severe outcomes.

**Results:** In 213,175 SARS-CoV-2 tests, despite similar positivity rates (8.2%F vs 8.9%M), males were more frequently hospitalized (28%F vs 33%M). Of 2,626 hospitalized individuals, females had less severe presenting respiratory parameters and males had more fever. Comorbidity burden was similar, but with differences in specific conditions. Medications relevant for SARS-CoV-2 were used at similar frequency except tocilizumab (M>F). Males had higher inflammatory lab values. In a logistic regression model, male sex was associated with a higher risk of severe outcomes at 24 hours (odds ratio (OR) 3.01, 95%CI 1.75, 5.18) and at peak status (OR 2.58, 95%CI 1.78,3.74) among 18-49 year-olds. Block-wise addition of potential explanatory variables demonstrated that only the inflammatory labs substantially modified the OR associated with male sex across all ages.

**Conclusion:** Higher levels of clinical inflammatory labs are the only features that are associated with the heightened risk of severe outcomes and death for males in COVID-19.

**Trial registration:** NA

**Funding:** Hopkins inHealth; COVID-19 Administrative Supplement (HHS Region 3 Treatment Center), Office of the ASPR; NIH/NCI U54CA260492 (SK), NIH/NIA U54AG062333 (SK).

## INTRODUCTION

Heterogeneity in the outcomes of SARS-CoV-2 infection has been a hallmark of the COVID-19 pandemic. Early reports from Wuhan, China^1,2^ and European countries^3^ showed heightened hospitalization, intensive care unit (ICU) admission, and mortality in males. Ongoing real-time surveillance^4^ and meta-analyses of over 3 million cases of COVID-19^5^ continue to reveal that while the proportion of COVID-19 cases are similar between the sexes, adult males are almost 3-times more likely to be admitted into ICUs and twice as likely to die as females. The reasons for this difference in outcomes remain poorly defined although multiple contributors including gender and sex-based factors have been proposed^6^.

Although analyses of global data provide little evidence for differences in exposure to SARS-CoV-2 between males and females, on a local level there are numerous cases of imbalanced exposure^5,7^. Differential exposure to SARS-CoV-2 is likely associated with behaviors, occupations, and societal and cultural norms (i.e., gender differences) that also impact the probability of access to testing and healthcare^8^. There are also gendered associations with health behaviors, including higher tobacco and alcohol use among males^9,10^, that have potential implications for COVID-19 pathogenesis^11,12^. The cross-cultural emergence of a male excess of severe outcomes suggests that while gender may impact exposure and health behaviors, the direct impact of biological sex differences in susceptibility to SARS-CoV-2 infection and pathogenesis of disease must also be considered^6^.

Sex is a modifier of the response to a number of viruses^13,14^, including influenza (higher fatality during reproductive age in females)^15^, HIV (lower HIV RNA levels, but the same rate of disease progression among females)^16^, hepatitis C (higher probability of spontaneous clearance among females)^17^, and SARS-CoV (males have a higher fatality rate than females)^18,19^. Immune responses are directly modified by sex steroids^20^ and sex-specific patterns of both autosomal and sex chromosome encoded genes^21^, which collectively contribute to sex differences in disease outcomes^13^. Mutations in the X-linked gene *TLR7* have sex-specific effects on susceptibility to severe COVID-19^22^. There also are differences in proinflammatory cytokine production (e.g., IL-6^23^) and T cell responses^24^ between males and females. In particular, although male and female COVID-19 patients have similar frequencies of virus-specific CD8+ T cells during infection, females maintain greater activity of CD8+ T cells, including proliferation, terminal differentiation, and IFN production, than males^24^. In contrast SARS-CoV-2 antigen specific IgM, IgG, and IgA, and neutralizing antibodies against whole virus, are consistently greater in male than female COVID-19 patients^25^ but the durability of neutralizing antibody titers over a six month period is reduced for males compared with females^26^. The significance of the sex differences in immune response in determining the outcome of infection are unknown, and few large studies have analyzed immune markers in the context of both sex and disease severity.

Worldwide, the case fatality rates (CFR) from COVID-19 increase with advancing age in both males and females^27^. However, the intersection of age and sex is complex. There are features of aging shared between males and females including increasing inflammation, immunosenescence, comorbidities, and risk of secondary infections all of which may contribute to age-related susceptibility to severe COVID-19 outcomes^27^. However, there are also sex specific features of aging, specifically the declining concentrations of sex steroid hormones, with specific points of shift around the menopausal transition among females. In the literature to date, all males, even the elderly are more likely to be hospitalized and die from COVID-19 than females^3,6,28^, but the magnitude of difference between males and females across age is less clear. Age-specific differences in the risk associated with male sex may provide important mechanistic insights into the role of sex steroid hormones.

Despite the many potential mechanisms and data demonstrating differences in immune phenotypes between males and females with COVID-19, few large studies have analyzed markers in the context of sex, age and disease severity and sought to link these responses to outcomes. Using data from over 200,000 SARS-CoV-2 tests and detailed clinical and demographic data from more than 2,600 individuals hospitalized with COVID-19 at our institution, we defined male-female differences in testing, admission, baseline comorbidities and health behaviors, medication use, laboratory markers, and outcomes. Our data identified imbalances in several factors, but point to a sex-specific profile of inflammation as the primary underlying driver of sex-differential outcomes, which is most prominent among individuals 18-49 years of age.

## METHODS

### Study design and participants

We performed a retrospective cohort analysis of patients who were tested for SARS COV-2 and treated between March 11, 2020 and October 31, 2020 at the Johns Hopkins Medicine health care system locations in the Maryland and Washington DC region. The study design and inclusion criteria for sequentially admitted patients with SARS-CoV-2 have been in part been described elsewhere^29^. This health system has 2,513 beds including 354 intensive care unit (ICU) beds and serves a population of approximately 7 million. It includes a network of referring clinics and 5 hospitals (Johns Hopkins Hospital, Baltimore, Maryland; Bayview Hospital, Baltimore, Maryland; Howard County General Hospital, Columbia, Maryland; Suburban Hospital, Bethesda, Maryland; and Sibley Hospital, Washington, DC). The data were organized as part the JH-CROWN registry: The COVID-19 PMAP Registry, which utilizes the Johns Hopkins Precision Medicine Analytics Platform to extract electronic health records^30^ and includes demographic characteristics, medical history, comorbid conditions, symptoms, vital signs, respiratory events, medications, and laboratory results.

The institutional review boards of these hospitals approved this study as minimal risk and waived consent requirements.

### Definitions and Outcome Measures

We stratified data by sex and with the following age categories: 18-49 (reproductive age), 50-64, 65-74 and 75 years or older. For testing data, we reported raw numbers and percentages for those who had a SARS-CoV-2 test result in the health system and included only the first positive test for any individual. Asymptomatic testing was performed in a number of scenarios in the hospital system including admission screening, screening on admission to labor and delivery, pre-procedural screening, surveillance protocols, post-exposure screening, and other miscellaneous indications (i.e. travel). Both asymptomatic and symptomatic testing was included for analysis. Test data by race and ethnicity is reported as the 7-day moving average of testing with definitions of Black or African American (non-Hispanic), White or Caucasian (non-Hispanic), Hispanic or Latino, or Other. For the admitted cohort, natural language processing was used to identify presenting symptoms, validated by manual abstraction on a subset of records as described previously^29^. Laboratory testing was determined by treating physicians. We report initial laboratory values as the median over the period of 48 hours prior to admission until 48 hours after time of admission, allowing inclusion of labs from outpatient visits immediately preceding presentation or transfer to the hospital of admission. Peak and nadir values are derived from all lab values during the course of the admission. For the analysis reported here, we focused on laboratory values postulated to have importance in SARS-CoV-2 including D-Dimer (coagulation), a set of inflammatory labs including ferritin, C-reactive protein (CRP), and IL-6, and the absolute lymphocyte count (ALC) and neutrophil:lymphocyte ratio (NLR) as lymphopenia has been a feature of COVID-19. Additional labs reflective of baseline health status in models included initial albumin, hemoglobin, estimated glomerular filtration rate (eGFR), and alanine aminotransferase (ALT).

Primary outcomes were defined by using the World Health Organization (WHO) COVID-19 disease severity scale^31^. The WHO scale is an 8-point ordinal scale ranging from ambulatory (1 = asymptomatic, 2 = mild limitation in activity) to hospitalized with mild to moderate disease (3 = room air, 4 = nasal cannula or facemask oxygen), hospitalized with severe disease (5 = high-flow nasal cannula or noninvasive positive pressure ventilation, 6 = intubation and mechanical ventilation, 7 = intubation and mechanical ventilation and other signs of organ failure [hemodialysis, vasopressors, extracorporeal membrane oxygenation]), and death (score of 8). We defined mild/moderate disease as a score of 3-4, severe disease as a score of 5 to 7, and the composite outcome of severe disease or death as a score of 5 to 8 on the WHO scale. Multiple comorbid condition burden was assessed by using the 17-item modified Charlson Comorbidity Index (CCI). Individual comorbidities were extracted from the medical records.

Medications were recorded as used if there was an order and at least one instance of a recorded time of administration. A treatment guidance document including clinical and laboratory features was used to inform the use of tocilizumab. All patients who met criteria were not offered the therapy and in rare cases (n=3) individuals who did not meet criteria were also given tocilizumab, but the majority of use was consistent with the guidance. These criteria included an IL-6 level >80 pg/mL or all of the following: D-dimer level >1 μg/mL, CRP level ≥10 mg/dL, Ferritin level >750 ng/mL in addition to oxygen requirement qualifications. We identified individuals with an SpO2:FiO2 (oxygen saturation:fraction of inspired oxygen) ratio of <256 as qualified for tocilizumab and determined total numbers of females and males who had qualifying values at any point during hospitalization.

### Statistical Analyses

We report the raw numbers and percentages of SARS-CoV-2 positive tests and hospital admission among persons with positive tests and the results of Chi-square tests to estimate differences by sex. Cohort characteristics were compared using Chi-square or Fisher’s exact test for small counts as noted in the table and figure legends. Continuous variables were compared with the t-test or, for non-Gaussian distributions, with the Wilcoxon rank sum test as indicated in the legends.

We developed a logistic regression model to estimate age-specific odds ratios comparing male and female incidence of severe disease/death versus mild/moderate disease among persons hospitalized. Final disposition status was available for all except for one of the individuals included in the cohort (>99.9%) at the time of analysis. The base model included hospital of admission and race/ethnicity and was stratified by age. We separately modeled two outcome variables: severe disease/death at 24 hours after admission (“24-hour” model); and at the most severe point during the hospital admission (“peak status” model). To determine the effect of potential mediating variables on the age-specific sex odds ratios, we divided variables of interest into 6 blocks. Each block was separately added to the baseline model to assess for modification of the sex effect in each age stratum in both the 24 hour and peak status model. The blocks are defined as follows: Block 1: BMI and admission source (nursing home versus other); Block 2: comorbidities including asthma, hypertension (complicated and uncomplicated), diabetes, chronic kidney disease, cardiovascular disease, COPD, and immune suppression; Block 3: health behaviors including smoking status -current, former, never, and alcohol use; Block 4: presenting vital signs including respiratory rate (RR), fever, SpO2:FiO2 ratio, pulse (median over the first 24 hours of presentation); Block 5: general status labs on presentation: albumin, ALT, hemoglobin, eGFR; Block 6: inflammatory labs: median initial value of ferritin, CRP, D-Dimer, ALC, NLR. Missing values for potential mediating variables were imputed prior to model fitting by using Multiple Imputation by Chain Equations (MICE).^32^ CRP, ferritin, ALC and ALT were log-transformed in the models. IL-6 was not included in the model because >50% of values were missing. We report point estimates with 95% confidence intervals for the age-specific sex effects and the block adjusted age-specific sex effects. We quantified the change in the age-specific sex odds ratios associated with block addition by reporting the change in the log odds ratio with a 95% confidence interval determined by bootstrapping. Sensitivity analyses of findings were performed by re-running the models excluding (1) pregnant women and (2) individuals with DNR/DNI orders within 24 hours of admission. All analyses were performed using R, version 4.0.2^33^.

## RESULTS

### SARS -CoV-2 testing and demographics

Between March and October 2020, there were 213,175 SARS-CoV-2 tests performed in the JHM system in individuals without a prior positive test result. Of those tests performed, 57% of tests were done in females and 43% in males. The overall test positivity rate was 8% for females and 9% for males, with females accounting for 55% of positive test results. More females tested positive in the <18 and 75 years and older age strata, and more males in all other age strata (**Figure 1A**). Females accounted for 43% of people with diagnosed SARS-CoV-2 infection who were admitted to the hospital. Within sex, the proportion of individuals with positive results who required hospital admission was higher in males compared to females between the ages of 18 and 74 years (**Figure 1B**). Of the tests performed, 102,760 (48%) were done on asymptomatic individuals (56% female versus 44% male tested). Indications for testing of asymptomatic individuals included: hospital admission screening (i.e. no COVID-19 symptoms but being admitted to the hospital for other reasons), COVID exposure history, pre-procedure testing, admission to labor floor with anticipated delivery, a surveillance screening program testing, and other (e.g., travel, imaging findings, or other required testing). Test positivity was similar, and slightly more frequent in males than females across all categories (overall test positivity rate 1.4% male, 1.2% female, p=0.05) including in the category of admission testing (n=21,915 tests) females were less likely to test positive (1.3% versus 1.6% of males, p=0.03) (**Figure 1C**). Proportion of positive tests for males and females tracked closely together within race/ethnicity groups (**Figure 1D**).

**Figure 1.**
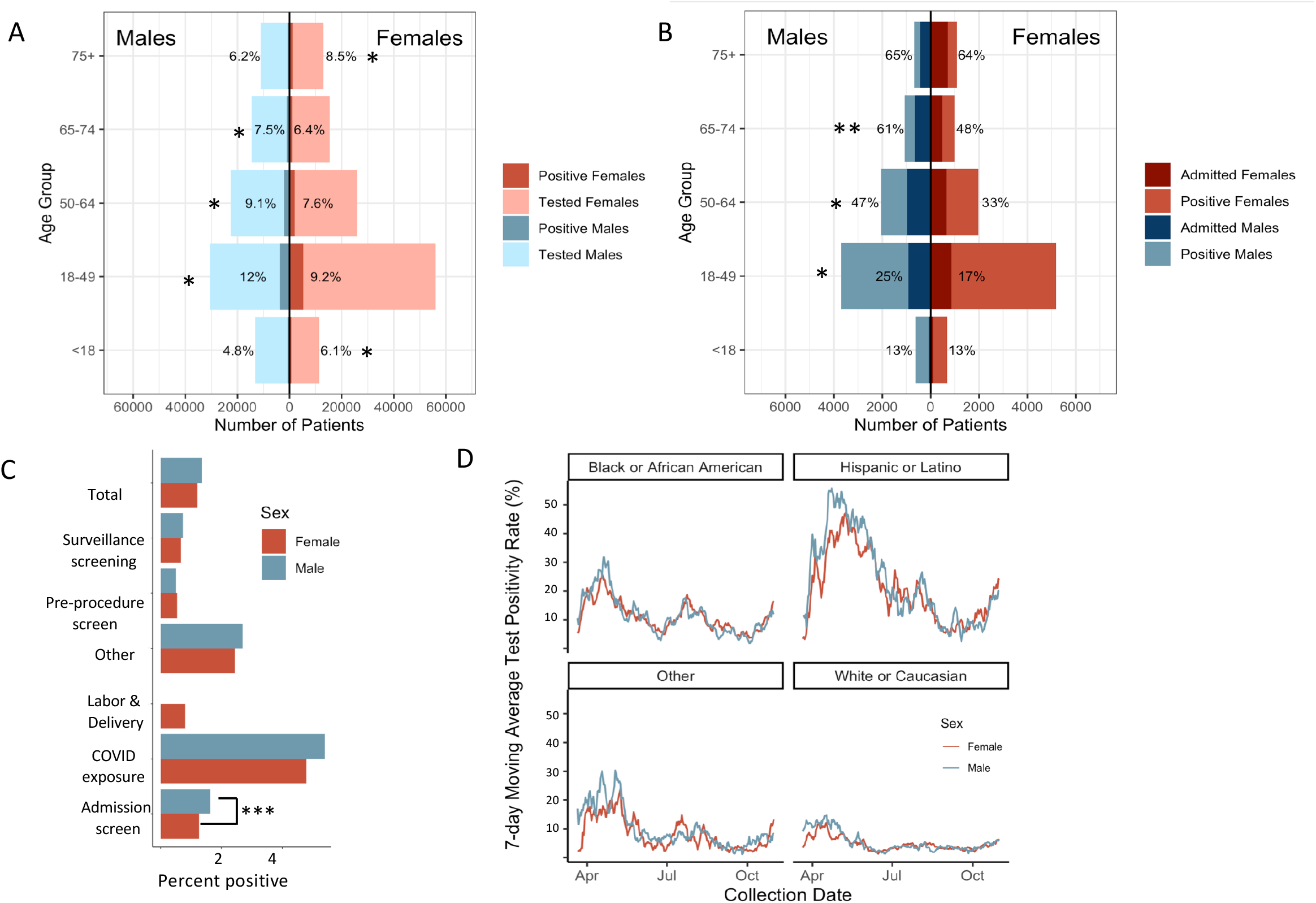
Testing and admission by sex and age. (A) number of COVID-19 tests and proportion positive by age group and (B) proportion of those testing positive who required hospital admission. *p<0.0001, **p<0.001, by Chi-square, asterisk is on the side with the higher value. Test positivity rates by sex among individuals considered asymptomatic at the time of testing (C) **p=0.03, by Chi-square. 7 day moving average of test positivity rates for males and females in different race/ethnicity groups (D).

### Clinical presentation and baseline comorbidities among those who were hospitalized

We analyzed the detailed clinical and demographic data for 2,626 patients admitted with SARS-CoV-2 infection during the study period (**Table 1**). Overall, males and females had similar median age but different distribution across age groups; females were enriched in the 75 years and older age stratum. Admission from a nursing home, which was previously shown to be associated with severe outcomes in this cohort^29^, was similar in frequency between males and females. Insurance status was distributed across multiple payor types in both males and females and was notable for more males in the “Other” category (males 17% versus females 11%, p<0.001) (i.e., insurance from worker’s compensation, Tricare, and other governmental programs). A greater proportion of females was Black and a greater proportion of males was White. A higher proportion of males had severe disease (males 36% versus females 28%, p<0.0001). Length of stay was approximately 0.5 days longer in males (p=0.02), and time to severe or death outcome was approximately 5 hours longer in females (p=0.04). Of note, there were numerically more females with DNR/DNI status within 24 hours of presentation, primarily driven by patients who were 75 years of age and older.

**Table 1.** Admission cohort characteristics stratified by sex. ^#^Admission source and race/ethnicity was available for >99% of cohort. ^&^Insurance source was not determined for ∼30% of the cohort. Time to outcomes presented in days with median and IQR shown. Statistics are Chi-square, except for^ which were compared by Wilcoxon rank sum test.

|  | **Females**<br>**N=1,280** | **Males**<br>**N=1,346** | **P value** |
| --- | --- | --- | --- |
| Median age, years (IQR)^ | 60 (42, 76) | 59 (46, 71) | 0.2 |
| Age distribution in strata, years |  |  |  |
| 18-49 | 431 (34%) | 413 (31%) | 0.1 |
| 50-64 | 300 (23%) | 420 (31%) | <0.0001 |
| 65-74 | 207 (16%) | 269 (20%) | 0.01 |
| >75 | 342 (27%) | 244 (18%) | <0.0001 |
| Admission from nursing home# | 193 (15%) | 191 (14%) | 0.6 |
| Insurance source& |  |  | 0.002 |
| Medicaid | 130 (15%) | 120 (14%) | 0.4 |
| Medicare | 368 (42%) | 341 (38%) | 0.09 |
| None | 49 (5.7%) | 59 (6.7%) | 0.4 |
| Other | 91 (11%) | 151 (17%) | <0.001 |
| Private | 228 (26%) | 216 (24%) | 0.4 |
| Race/ethnicity# |  |  | 0.002 |
| Black | 504 (40%) | 463 (34%) | <0.01 |
| White | 299 (23%) | 400 (30%) | <0.001 |
| Hispanic | 128 (10%) | 130 (9.7%) | 0.8 |
| Other | 342 (27%) | 350 (26%) | 0.7 |
| Median BMI (IQR)^ | 30 (25, 36) | 28 (24, 32) | <0.001 |
| Severe disease/death | 358 (28%) | 482(36%) | <0.0001 |
| Ventilated | 69 (5.5%) | 107 (8.0%) | 0.01 |
| Death | 149 (12%) | 182 (14%) | 0.2 |
| Time to severe/death outcome, days (IQR)^ | 0.84 (0.06, 3.05) | 0.63 (0.02, 2.76) | 0.04 |
| Length of stay, days (IQR)^ | 5.5 (2.6, 10.7) | 6.0 (2.9, 11.6) | 0.02 |
| DNR/DNI within 24h | 80 (6.2%) | 61 (4.5%) | 0.06 |

Overall, symptoms reported on presentation were similar between the sexes, but a greater proportion of males reported fevers (p<0.05), whereas females had a greater frequency of headache (p<0.001), loss of smell (p<0.05), and vomiting (p<0.001) (**Figure 2**). When symptom data were disaggregated into age strata, the loss of smell was primarily driven by a significant difference in the 18-49 years age group (33% of females versus 25% of males in that age group, p=0.01). This age group also uniquely showed a difference in frequency of cough reported, with males reporting cough more frequently than females (86% males versus 78% females, p<0.01).

**Figure 2.**
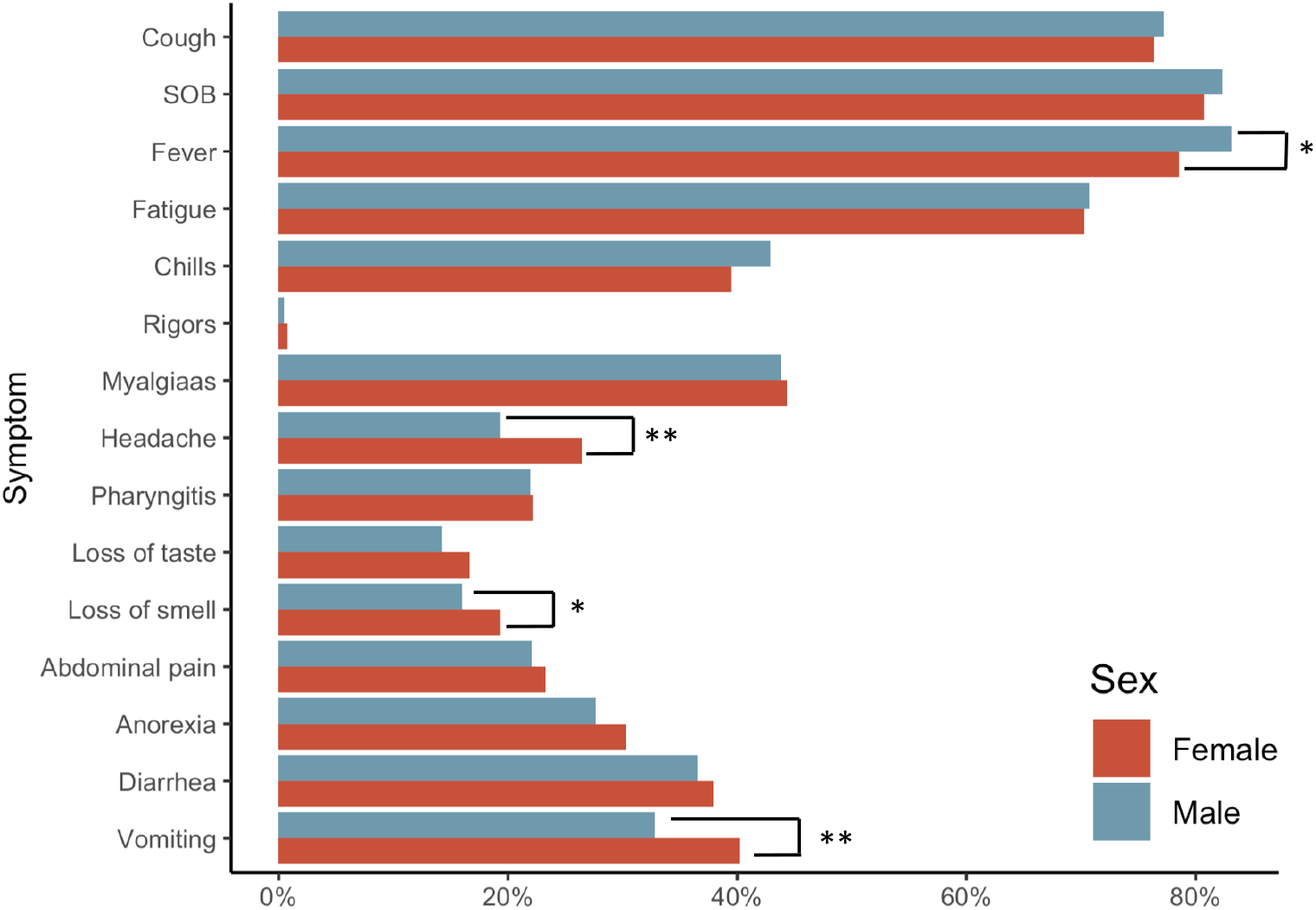
Distribution of reported symptoms for males and females. Natural language processing was used to extract symptom features from chart notes with complete data available for approximately 91% of the cohort. Vomiting, headache, and loss of smell were more common in females, and fevers were more commonly reported in males. *p<0.05, **p<0.001, by Chi-square.

Vital signs on presentation were consistent with the symptoms reported. More males than females had a measured temperature > 38.0°C on presentation (p<0.001). In contrast, females presented with more favorable respiratory parameters, including lower respiratory rates (RR), lower levels of supplemental O2, and a greater SpO2:FiO2 ratio across all age groups (p<0.001) (**Table 2**). In the subset of individuals who developed severe disease or progressed to death (n=843, 32%), the sex differences in initial presenting vital signs were attenuated but persisted for the proportion of males with a recorded temperature >38.1 (p<0.001). (**Table 2**). At both six and 24 hours after admission, WHO status was significantly different by sex (**Supplemental Table 1**), with females more frequently classified as mild/moderate than males.

**Table 2.** Signs on presentation. Vital signs from the first 24 hours after hospital admission among all individuals and those who developed severe disease or progressed to death. For temperature, shown are number febrile (%), for all other values, median (standard deviation) over the first 24 hours are shown. Statistics for categorical variables are by Chi-square, and by t-test for continuous variables.

|  | Overall cohort |  |  | Severe/death cohort |  |  |
| --- | --- | --- | --- | --- | --- | --- |
|  | All ages |  |  | All ages |  |  |
|  | Females<br>N=1280 | Males<br>N=1346 | P value | Females<br>N=360 | Males<br>N=483 | P value |
| Temp >38.1 °C | 383 (31%) | 566 (42%) | <0.001 | 141 (39%) | 263 (55%) | <0.001 |
| Pulse beats/<br>minute | 85 (15) | 86 (15) | >0.9 | 88 (17) | 89 (17) | 0.05 |
| Respiratory rate<br>breaths/minute | 20.4 (4.3) | 21.1 (5.0) | <0.001 | 23.2 (5.9) | 24.1 (6.2) | 0.03 |
| FiO<sub>2</sub> | 0.30 (0.17) | 0.33 (0.20) | <0.001 | 0.45 (0.25) | 0.49 (0.26) | 0.03 |
| SpO<sub>2</sub>:FiO<sub>2</sub> ratio | 384 (119) | 364 (130) | <0.001 | 275 (129) | 257 (132) | 0.05 |

Abbreviations: Temp: temperature; FiO<sub>2</sub>: fraction of inspired oxygen; SpO<sub>2</sub>: oxygen saturation.

The overall burden of comorbid conditions was similar between males and females as measured by the Charlson comorbidity scores (**Table 3**). However, significant differences in specific comorbid conditions were observed including a higher prevalence of chronic lung disease and asthma among females, in particular younger adult females (p<0.001) (**Table 3, Figure 3A**). This diagnosis code was validated by a cross check of admission medications which indicated 95% of those with an asthma diagnosis had an asthma medication as part of their outpatient regimen. Obesity also showed a sex and age intersection, with higher BMI values for females in all age strata, in particular in the <75 age group (**Table 3, Figure 3B**). Chronic kidney disease and complicated hypertension were more common among males (**Table 3, Figure 3A)**. Males also had a higher frequency of both smoking and alcohol use (**Table 3, Figure 3C,D**).

**Table 3.** Baseline comorbid conditions and health behaviors by sex. Statistics by Chi-square except for Charlson score which was done by t-test. Abbreviations: BMI: Body mass index.

|  | Females<br>N=1,269 | Males<br>N=1,339 | P value |
| --- | --- | --- | --- |
| BMI category* |  |  | <0.001 |
| <30 | 505 (47%) | 707 (63%) |  |
| 30-35 | 243 (23%) | 235 (21%) |  |
| 35-40 | 156 (15%) | 108 (9.6%) |  |
| >40 | 163 (15%) | 79 (7.0%) |  |
| Comorbidity burden |  |  |  |
| Charlson score<br>[Mean(S.D.)] | 1.8 (1.8) | 1.9 (1.8) | 0.7 |
| Comorbid conditions |  |  |  |
| Congestive heart failure | 241 (19%) | 276 (21%) | 0.3 |
| Cardiovascular disease | 489 (38%) | 560 (42%) | 0.08 |
| Hypertension |  |  |  |
| Complicated | 291 (23%) | 376 (28%) | 0.003 |
| Uncomplicated | 682 (53%) | 725 (54%) | 0.8 |
| Liver disease | 143 (11%) | 183 (14%) | 0.07 |
| Asthma | 231 (18%) | 114 (8.5%) | <0.001 |
| Chronic pulmonary disease | 389 (30%) | 287 (21%) | <0.001 |
| Chronic kidney disease | 225 (18%) | 311 (23%) | <0.001 |
| Diabetes |  |  |  |
| Type I | 23 (1.8%) | 24 (1.8%) | >0.9 |
| Type II | 462 (36%) | 509 (38%) | 0.4 |
| Cancer | 140 (11%) | 138 (10%) | 0.7 |
| Transplant | 29 (2.3%) | 40 (3.0%) | 0.3 |
| HIV | 14 (1.1%) | 25 (1.9%) | 0.2 |
| Health behaviors |  |  |  |
| Alcohol use<sup>#</sup> | 209 (19%) | 391 (34%) | <0.001 |
| Smoking status<sup>%</sup> |  |  | <0.001 |
| Current Smoker | 44 (3.8%) | 112 (9.5%) |  |
| Former Smoker | 246 (21%) | 295 (25%) |  |
| Never Smoker | 868 (75%) | 773 (66%) |  |

**Figure 3.**
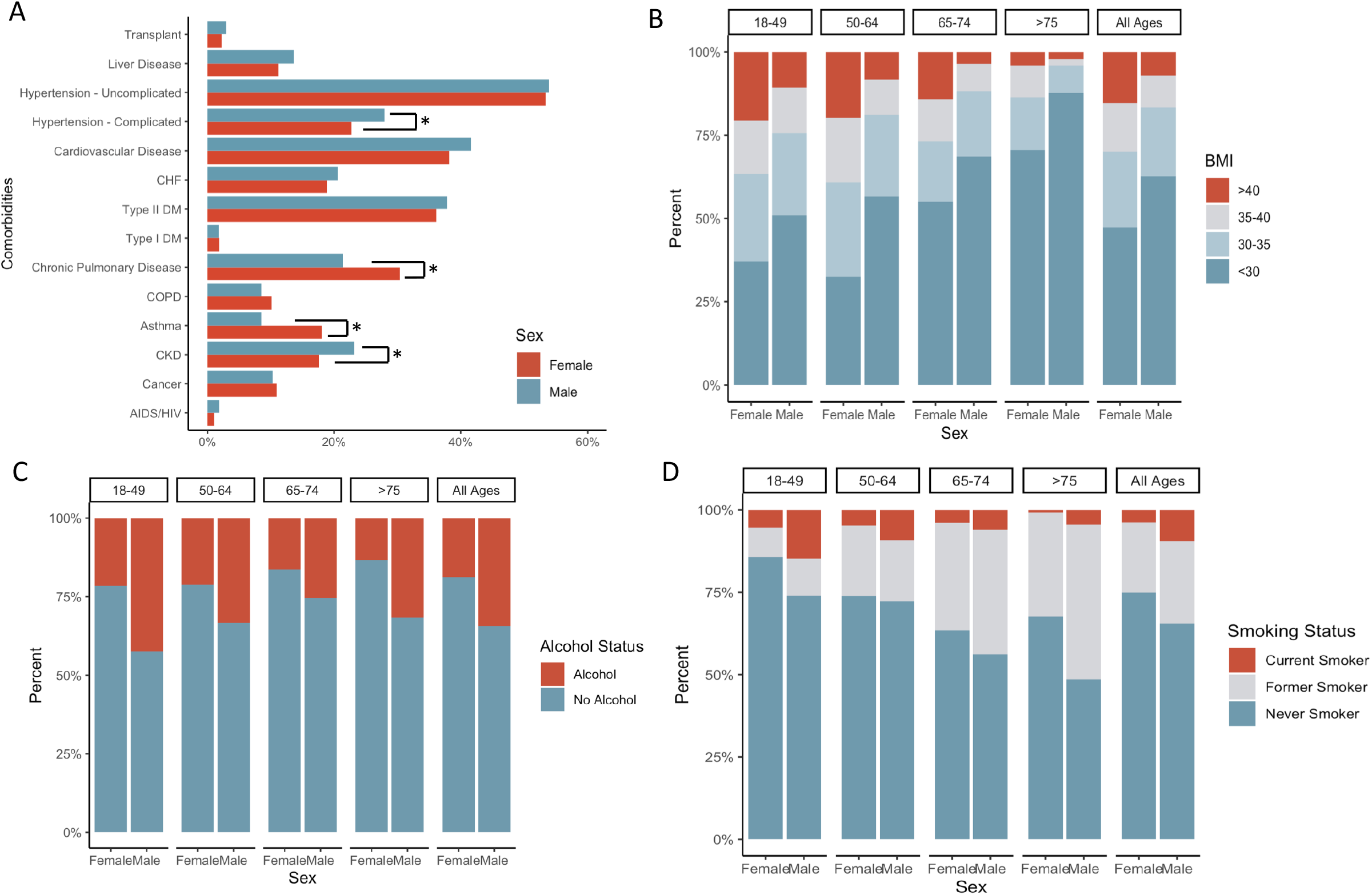
Frequency of comorbid conditions at baseline for males and females, with several comorbidities presenting with a sex imbalance. (A) (*p<0.05, **p<0.001, by Chi-square.) Distribution of BMI categories (B), alcohol use (C) and smoking status (D) by sex and age. (Overall Chi-square for B, C, and D, p<0.001).

### Laboratory measures

In the first 48 hours of admission, females and males had comparable levels of D-dimer and erythrocyte sedimentation rate (ESR), but males had lower absolute lymphocyte counts, higher neutrophil:lymphocyte ratios (NLR), and higher ferritin, IL-6 and CRP levels than females **(Figure 4**, p<0.001 Wilcoxon rank sum test). Age stratified analyses demonstrated differential effects of age on inflammatory markers; with increasing age, there was less difference between the median values of males and females for CRP, ferritin and IL-6. For IL-6, extremely elevated values were specifically more frequent among males in the 65-74 and 75-and-older age strata whereas the median separation was most notable for the 18-49 years age stratum (**Figure 4**). To assess whether this difference was related to timing in disease course and time of presentation, we assessed the peak (i.e., CRP, ferritin, IL-6, NLR) or nadir (i.e., ALC) levels over the course of the admission and observed the same patterns of lower markers of inflammation in females (**Supplemental Table 2)**.

**Figure 4.**
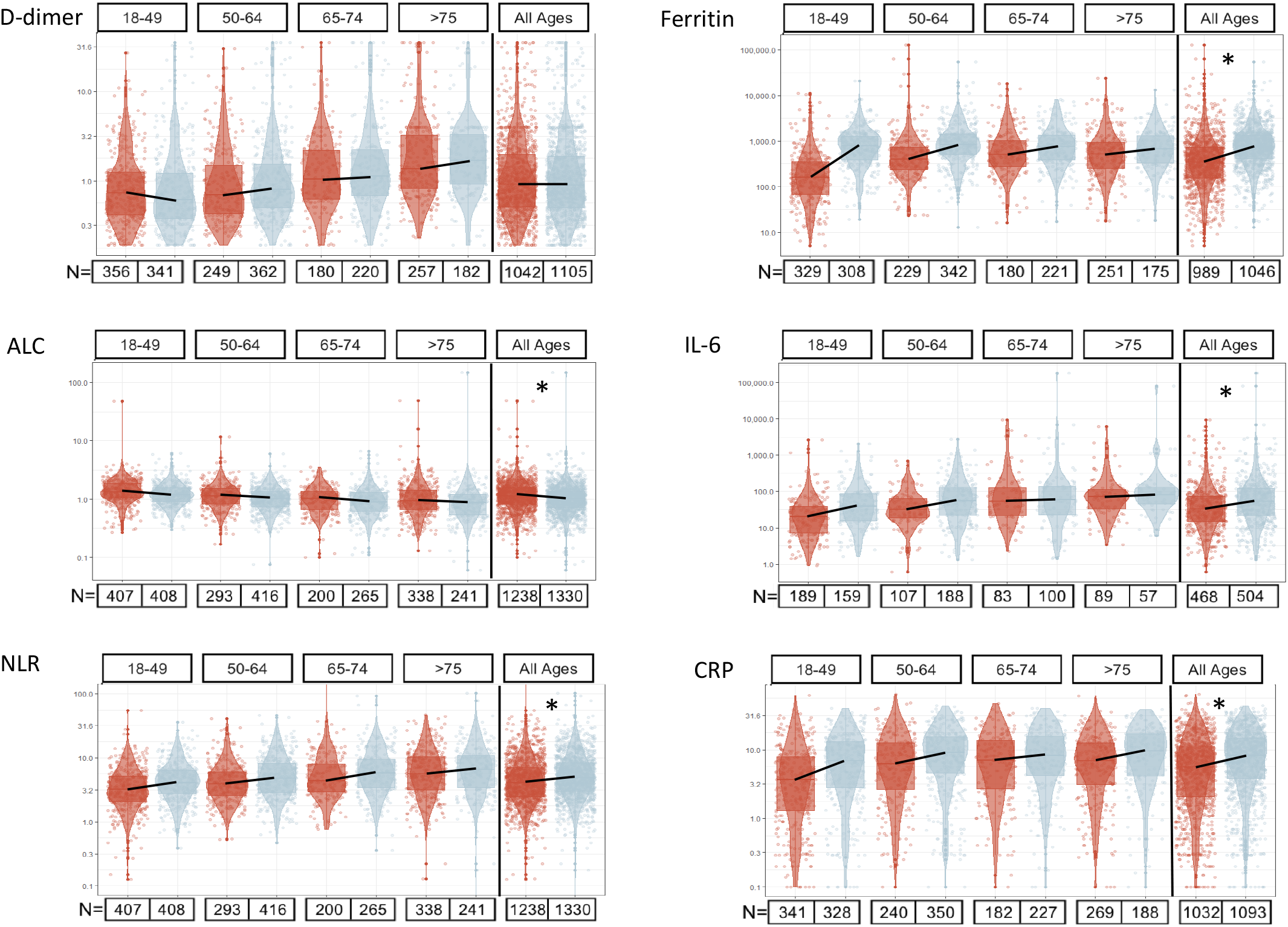
Median lab values at the time of admission (−48 hours to +48 hours) by sex and age. Red are female and blue are male, total numbers of observations for each lab are indicated below the column and the rightmost column is the overall sex comparison. Statistics on the overall comparison, *p<0.001 by Wilcoxon rank test. ALC=absolute lymphocyte count, NLR= neutrophil:lymphocyte ratio.

### Medication use

Using the detailed medical records, we identified use of medications postulated to be relevant to the course of SARS-CoV-2 infection. First, we assessed baseline use of angiotensin converting enzyme (ACE) inhibitors, angiotensin receptor blockers (ARBs), and statins among males and females in the cohort (**Table 4**). Baseline use of statins and ARBs was comparable between males and females, but males had a greater frequency of ACE inhibitor use (155 males (12%) versus 114 females (9%), p=0.03).

**Table 4.** Medication use at baseline and during the course of hospitalization for SARS-CoV-2 Infection. Statistics by Chi-square. Data are complete for >98% of cohort.

| Medication N(%) | Females<br>N=1,280 | Males<br>N=1,346 | P value |
| --- | --- | --- | --- |
| **Agents directed at COVID-19** |  |  |  |
| Hydroxychloroquine | 187 (15%) | 225 (17%) | 0.2 |
| Remdesivir | 194 (15%) | 235 (18%) | 0.1 |
| Tocilizumab | 20 (1.6%) | 74 (5.6%) | <0.0001 |
| Corticosteroids | 324 (26%) | 317 (24%) | 0.3 |
| **Baseline medications with potential relevance to COVID-19 pathogenesis** |  |  |  |
| ACE inhibitors | 114 (9.0%) | 155 (12%) | 0.03 |
| ARBs | 105 (8.3%) | 100 (7.5%) | 0.5 |
| Statins | 365 (29%) | 425 (32%) | 0.1 |
| **Agents directed at coinfections** |  |  |  |
| Antibiotics | 812 (64%) | 884 (66%) | 0.3 |
| Antifungals | 108 (8.5%) | 97 (7.3%) | 0.3 |

Analysis of those medications that were intended as therapeutics against COVID-19 revealed similar patterns of usage between males and females for remdesivir, hydroxychloroquine, and steroids (**Table 4**). Of note, despite the higher frequency of asthma and chronic lung disease among females we did not see an excess of steroid use during hospitalization in the females. The use of tocilizumab, however, was more common in males than females (p<0.0001). We also assessed the frequency of qualifying for tocilizumab use, based on laboratory values and hypoxemia outlined in the local treatment guidance. Females less frequently met these criteria, but even among those who did meet laboratory criteria and were hypoxemic, females received tocilizumab less frequently than males (8.3% of females and 20% of males, p <0.001) (**Table 5**).

**Table 5.** Number of females (F) and males (M) who qualified for tocilizumab use based on hypoxemia and inflammatory laboratory values as specified in the text and the number of those who qualified who received therapy. Frequency of use among those who qualified for consideration was significantly higher among males than females (data shown age stratified, statistics by Chi-square comparing females and males of all ages).

|  | 18-49 yrs |  | 50-64 yrs |  | 65-74 yrs |  | ≥75 yrs |  | Overall |  | P |
| --- | --- | --- | --- | --- | --- | --- | --- | --- | --- | --- | --- |
| Tocilizumab | F=431 | M=413 | F=300 | M=421 | F=207 | M=269 | F=342 | M=244 | F=1280 | M=1346 | value |
| Qualified | 20 | 73 | 50 | 126 | 64 | 90 | 71 | 66 | 205 | 355 | **<0.001** |
| Received | 2 | 15 | 4 | 25 | 6 | 20 | 5 | 12 | 17 | 72 |  |

### Modeling the impact of sex on the risk of severe disease

To assess the impact of sex on the risk for severe COVID-19 or death, we developed an age stratified logistic regression model including the baseline demographic variables of race/ethnicity and hospital of initial admission. We first tested the impact of sex on early mortality (i.e., within 24 hours of presentation), and found the OR (all shown with [95% CI]) for severe disease or death at 24 hours for males relative to females were 3.01 [1.75,5.18] in the 18-49 years age group, 1.41 [0.95,2.10] in the 50-64 years age group, 1.47 [0.92,2.35] for individuals 65-74 years of age, and 0.77 [0.48,1.24] in patients >75 years of age (**Figure 5**). We then used a blockwise addition approach to test the impact of other variables on the male excess in severe COVID-19 and death. Each block addition (black filled symbol) is shown in **Figure 5** in reference to the baseline model (i.e., the gray open symbols). With the addition of blocks for BMI/admission source, comorbidities, and health behaviors (i.e., smoking and alcohol use), there was minimal change in the odds ratio of severe COVID-19 outcomes. Adding the presenting vital signs did shift the estimated sex effect, consistent with the known predictive value of respiratory parameters and hypoxemia in severe outcomes. Baseline laboratory values, used to indicate general health status, had a modest impact on the sex risk for severe outcomes. Addition of the block of inflammatory laboratory values, however, substantially reduced the odds ratio of risk of severe outcomes for males, most prominently in the 18-49 age group (**Figure 5**). To quantify the impact of block addition, we assessed the change in log OR with confidence intervals for each block addition. Across all the ages, addition of the inflammatory labs was the only block that significantly changed the estimated sex effect (p<0.0001, **Supplemental Figure 1**).

**Figure 5.**
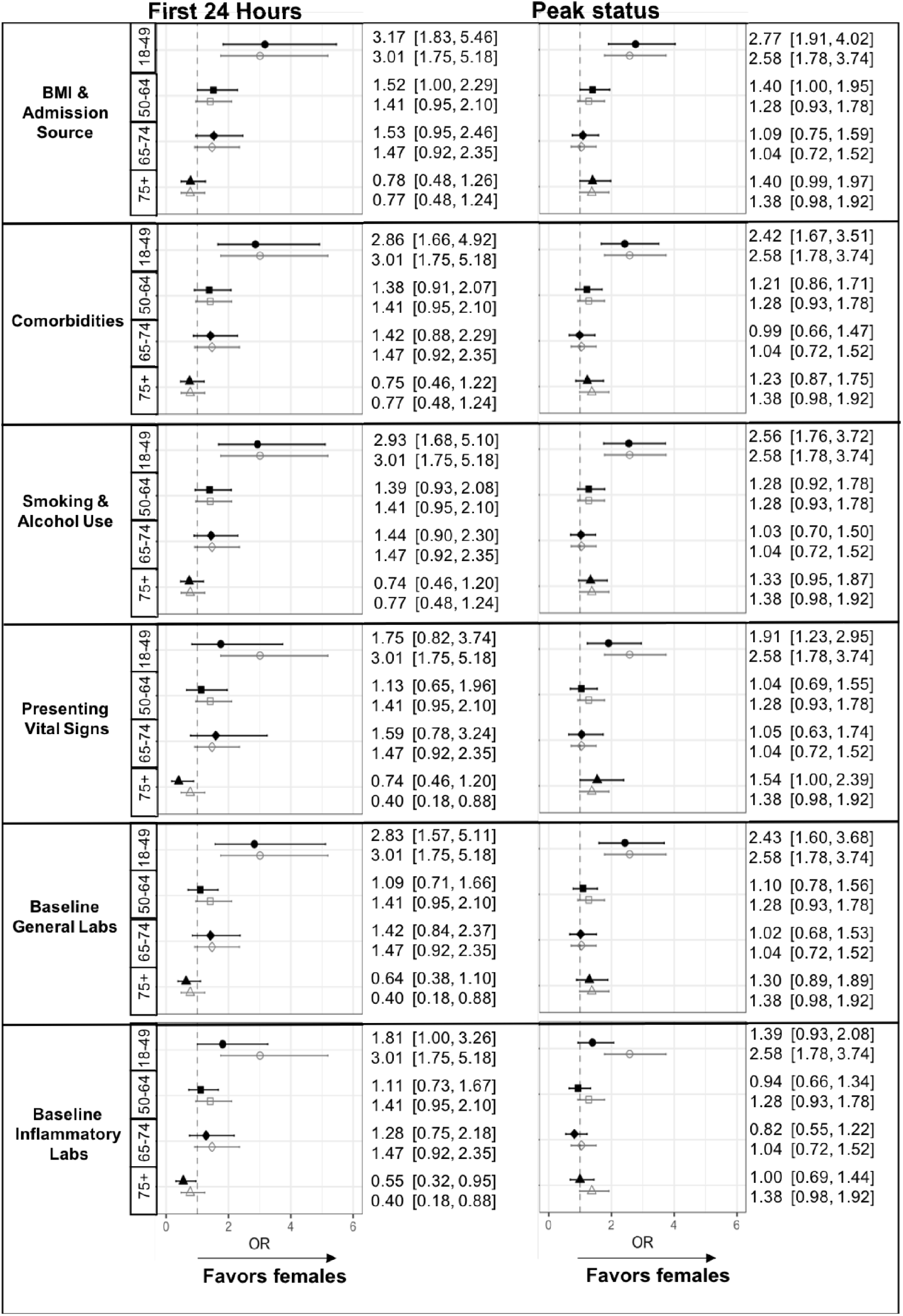
Odds ratio of increased risk for severe or death outcomes in males stratified by age. The risk of male sex is represented for the baseline model (open gray symbol in each graph) and then after addition of each block of variables (black filled symbol) for a model of outcomes at 24 hours and at peak status during the hospitalization. The numbers indicate the point estimate for the OR and the 95% CI.

The same analysis was repeated to assess sex differences in peak disease status at any point during the hospitalization. The OR of severe or death outcomes was significantly elevated in males between the ages of 18-49 years with an OR of 2.58 [95% CI 1.78,3.74] (**Figure 5**). Blockwise addition of variables again showed minimal change with the blocks of comorbidities, health behaviors, and baseline labs. The presenting vital signs still modified the OR of peak status, although somewhat more modestly than in the 24-hour model. Again, the initial inflammatory labs substantially shifted the OR for severe outcomes associated with male sex in particular in the 18-49 year old age stratum (sex effect from 2.58 [95% CI, 1.78,3.74] baseline model to 1.39, [95% CI 0.93,2.08] after inflammatory lab block addition). Across all ages, the addition of the inflammatory lab block was the only block associated with a statistically significant change in the log adds (p<1e10^−14^, **Supplemental Figure 1**). In both models, the increased risk of severe COVID-19 outcomes in males was primarily in the 18-49 years old age stratum. Sensitivity analyses excluding all pregnant women and individuals with DNR/DNI status within 24 hours did not substantially change the analytic findings (data not shown).

## Discussion

In this study, we leverage the power of a large hospital system with detailed patient level data to elucidate features of COVID-19 infection that differ between males and females from diagnosis to outcomes. Our data are consistent with prior reports of more severe disease in males^3,28,34-37^, but our analysis adds novel insights into asymptomatic test positivity rates, differential features of hospital presentation, and identification of inflammatory markers as predictors of the sex differential risk of severe COVID-19 outcomes. These analyses also indicate that the largest sex effect is in the youngest age stratum (18-49 years) with potential implications for mechanisms of a female protective effect. Our data suggest that there is a fundamental difference in the immune inflammatory response to SARS-CoV-2 infection that is advantageous in females, in particular in females of reproductive age.

We assessed sex-distribution of testing to address potential differences in identification of infection that might impact the estimated sex effect on the risk of severe disease. Our results support comparable rates of test positivity but higher rates of hospitalization among males, consistent with prior data from France^3^, New York^38^, Houston ^35^, and New Orleans^37^. An unresolved question is whether the difference in disease severity extends to the level of asymptomatic infection. A prior study using contact-based surveillance versus symptom-based surveillance in case identification had greater rates of cases among females with contact-based testing, supporting the hypothesis that asymptomatic infections are more common among females^39^. Our data do not support this finding; in >100,000 tests of asymptomatic individuals, the rate of test positivity between males and females was comparable. It is notable that our results among asymptomatic individuals also differ from the results of a serotesting survey in a large cohort of volunteers (11,283 participants) without a known history of SARS-CoV-2 infection or recalled symptoms^40^. Seropositivity rates in that study were higher among females at 5.5% compared to 3.5% of males (182 versus 122 positive tests in females and males, respectively)40. This data is notable in the setting of other studies which report higher sensitivity of antibody testing in males versus females after documented SARS-CoV-2 infection^41^. In our study, asymptomatic cases were identified during infection with direct testing for SARS-CoV-2 RNA and not by antibody testing. There may be sex differences in nasal viral load, as lower viral loads or shorter windows of viral shedding would bias RNA-based testing against identifying infection in females. Further work is needed to explore these possibilities. Taken together, gender-based risks, including overrepresentation of females among healthcare workers^42,43^, differences in care-seeking behaviors^44^ and perception of risk and acceptance of mitigation strategies^45^ may all contribute to risk of SARS-CoV-2 infection. Our data do not suggest significant differences in asymptomatic infection, and have overall similar rates of positivity in males and females. Notable, the difference in severe outcomes persists despite an overrepresentation of older females in the admitted cohort.

Prior studies have also identified an increased risk of severe outcomes and death among males^28,36,38^. In a multinational study of medical record data, male mortality was greater, and despite differences in comorbidities, the risk of death remainder greater for males than females, even in models with propensity score matching^46^. It has been difficult to determine which factors underlie a protective effect among females, hence in our analysis, we sought to identify potential contributors to sex and gender differences. While our data does show different prevalence in several of the descriptive features of the cohort (e.g. elevated BMI, complicated hypertension, asthma, alcohol and tobacco use), to our surprise none of these variables significantly altered the estimated sex effect on the risk of severe disease. This is notable in light of the high prevalence of obesity among females in our cohort, as obesity has been associated with progression to severe disease in multiple analyses^29,36^. We also observed a minimal difference in time to peak disease status, arguing against a gender-based difference in the timing of seeking care. Our analytic approach of adding variables by block allowed us to broadly query potential sources of variation between males and females and clearly emphasize differences in inflammation as a critical contributor to outcomes.

We also assessed whether there were differences in therapeutic interventions targeting SARS-CoV-2 infection. In our cohort, females were less likely to receive the immunomodulatory therapy tocilizumab, even when they fulfilled the institutional criteria for considering use of this therapeutic. Multiple factors converge on the decision to use a therapeutic without proven efficacy, including clinician decision making, patient acceptance of treatments and the biomedical features of the disease and we cannot draw definitive conclusions about the role of sex/gender in these decisions. It is notable that the trend towards lower inflammatory markers reduced the number of female patients who met a threshold for consideration of this particular therapeutic. There has been little work done to identify whether there are differences in receipt or efficacy of therapies targeting COVID-19 disease. In the sex subgroup analysis of dexamethasone for COVID-19, females did not have a significant benefit from therapy, although this may reflect lower disease severity among females in the trial^47^. Likewise, in the recently released results from the RECOVERY trial tocilizumab arm, while they observed a benefit of treatment among patients with hypoxia and systemic inflammation, in subgroup analysis, females did not have a significant benefit from therapy^48^. Further work is needed to determine whether there are gender differences in the use of immunomodulatory treatments more broadly and careful analysis of clinical trial data may determine whether there are sex-specific inflammatory thresholds that could guide use of anti-inflammatory agents in COVID-19 patients.

In our examination of outcomes by sex, male sex was associated with an increased risk for severe disease or death in both models assessing first 24 hours post admission as well as peak illness during the course of hospitalization. Block-wise addition of factors including BMI, admission source, comorbidities, health behaviors, and baseline general status labs had a minimal impact on male-associated risk. The presenting vital signs impacted the sex effect, more for model assessing outcomes within the first 24 hours of admission than for the peak illness status model. What was most notable in our study was the influence that the inflammatory marker profile had on male sex risk of severe outcomes. In fact, it was the only block that was statistically significant in both the first 24 hours and peak illness models across the full cohort. These data are consistent with a differential inflammatory response to SARS-CoV-2 infection as a driver of increased male risk. The clinical indices used in these analyses (i.e., CRP, ferritin, ALC, NLR) are biomarkers and likely not direct mediators of disease outcomes. These findings suggest that further research should seek to identify the sex-specific immune response features that contribute to this inflammatory profile. Once such example is the severe disease phenotype in male patients with mutations in *TLR7*^22^, which encodes an RNA-sensing receptor upstream of interferon production encoded on the X chromosome with well-described differences between males and females^49,50^. Differences between males and females in the peripheral immune phenotype have also been described^24^, but it is not known how these differences arise. It remains unclear whether the sex effect is a shift in immune setpoint or a difference in pathway activation, answering that question could guide the clinical utility of immune therapies in both sexes.

In summary, our methodological approach of carefully assessing the effect of risk factor groups on the sex-specific risk for severe COVID-19 illness has identified that inflammation profiles and not expected sociodemographic and other clinical characteristics are what are most associated with adult males being significantly more likely than females to progress to severe COVID-19 illness. This finding is most notable in the 18-49 year age stratum.

Future mechanistic studies must consider whether sex steroid hormones, sex chromosome complement, autosomal gene expression differences, or a combination of factors drives the overall female advantage not only in COVID-19^6^ but for other respiratory infections as well^51^. Age stratification is necessary to address some of these questions, and the marked increase in male risk in the 18-49 years old age stratum supports the hypothesis that sex steroid hormones have a role in the female protective effect. The data across the life span must be considered carefully however, as the female survivorship advantage among older adults (75+) is weighed against the higher numbers of women in this age category and with an early DNR/DNI status order. The data presented here, however, support a greater need to focus on immune pathway differences in males and females of reproductive ages (i.e., 18-49 years).

## Data Availability

The data for this study is maintained in the Johns Hopkins Precision Medicine Analytics Platform (PMAP).

## Author contributions

EPS, SLK, AG conceived of the study, and together with GS, MF, GM, KBR, and SZ designed the statistical analysis plan. GS, MF, GM, JM, JB contributed to data extraction, cleaning, analysis and presentation, EK contributed testing and admission data. BTG, MR, JM, JB contributed to the creation and maintenance of the JH-CROWN Registry. All of the authors contributing to the writing and editing of the manuscript. EPS, GS, and MF had full access to all of the data and are responsible for the integrity of the data and accuracy of analysis.

## Acknowledgements

The data utilized were part of JH-CROWN: The COVID PMAP Registry, which is based on the contribution of many patients and clinicians and is funded by Hopkins inHealth, the Johns Hopkins Precision Medicine Program. Drs. Garibaldi, Muschelli, Robinson, Bandeen-Roche, and Gupta and Mr. Schumock received funding from the COVID-19 Administrative Supplement for the HHS Region 3 Treatment Center from the Office of the Assistant Secretary for Preparedness and Response. Drs S. Klein, Zeger and Mr. Betz received funding from the NIH/NCI funded COVID-19 Serology Center of Excellence (U54CA260492). Dr. Scully is supported by R01AI154541 (NIAID/ORWH). The funders had no role in the design, analysis, or conduct of the study or in the decision to submit the manuscript for publication. The authors would like to acknowledge the efforts of many people throughout the institution who have worked to focus resources on the response to the COVID-19 epidemic.

---

**Supplemental Table 1.** WHO category at 6 hours and 24 hours after admission stratified by age and sex. Cells are the absolute number of events at each time and the within-sex percentage of that status. Distribution of WHO status was significantly different by sex at both timepoints when all ages were considered together, p<0.001 by Fisher’s exact at 6 hours and p<0.001 at 24 hours by Chi-square. Age stratified data is shown to illustrate the numbers and distribution of status across the cohort. Abbreviations. Mod: moderate.

| Age group, years | WHO status at 6 hours |  |  |  |  |  | WHO status at 24 hours |  |  |  |  |  |
| --- | --- | --- | --- | --- | --- | --- | --- | --- | --- | --- | --- | --- |
|  | Females, n (%) |  |  | Males, n (%) |  |  | Females, n (%) |  |  | Males, n (%) |  |  |
|  | Mild/Mod | Severe | Death | Mild/Mod | Severe | Death | Mild/Mod | Severe | Death | Mild/Mod | Severe | Death |
| 18-49 | 414<br>(96%) | 17<br>(3.9%) | 0<br>(0%) | 370<br>(90%) | 43<br>(10%) | 0<br>(0%) | 407<br>(94%) | 23<br>(5.3%) | 1<br>(0.2%) | 348<br>(84%) | 64<br>(15%) | 1<br>(0.2%) |
| 50-64 | 267<br>(89%) | 33<br>(11%) | 0<br>(0%) | 352<br>(84%) | 67<br>(16%) | 1<br>(0.2%) | 244<br>(81%) | 54<br>(18%) | 2<br>(0.7%) | 316<br>(75%) | 101<br>(24%) | 3<br>(0.7%) |
| 65-74 | 177<br>(86%) | 29<br>(14%) | 1<br>(0.5%) | 218<br>(81%) | 51<br>(19%) | 0<br>(0%) | 159<br>(77%) | 46<br>(22%) | 2<br>(1.0%) | 196<br>(73%) | 65<br>(24%) | 8<br>(3.0%) |
| >75 | 301<br>(88%) | 38<br>(11%) | 3<br>(0.9%) | 216<br>(89%) | 26<br>(11%) | 2<br>(0.8%) | 279<br>(82%) | 50<br>(15%) | 13<br>(3.8%) | 203<br>(83%) | 34<br>(14%) | 7<br>(2.9%) |
| All ages | 1,159<br>(91%) | 117<br>(9.1%) | 44<br>(0.3%) | 1,156<br>(86%) | 187<br>(14%) | 3<br>(0.2%) | 1,089<br>(85%) | 173<br>(14%) | 18<br>(1.4%) | 1,063<br>(79%) | 264<br>(20%) | 19<br>(1.4%) |

**Supplemental Table 2.** Median initial laboratory values (from −48 to +48 hours around the time of admission) and the median of the maximum (or nadir for ALC) values for the entire hospital course shown by sex. Number of samples for each measure (n) is indicated below the median value. Statistics with Wilcoxon rank sum tests, shown are median and interquartile ranges for all values.

|  | Median initial values |  | P value | Peak or nadir values |  | P value |
| --- | --- | --- | --- | --- | --- | --- |
|  | Females<br>N=1,258 | Males<br>N=1,340 |  | Females<br>N=1,267 | Males<br>N=1,343 |  |
| Laboratory measure | Median (IQR) | Median (IQR) |  | Median (IQR) | Median (IQR) |  |
| Absolute lymphocyte count, $\times 10^3$ cells/μL | 1.1 (0.82, 1.6) | 1.0 (0.71, 1.4) | <0.001 | 0.87 (0.53, 1.3) | 0.74 (0.45, 1.1) | <0.001 |
|  | n=1238 | n=1330 |  | n=1255 | n=1339 |  |
| CRP, mg/dL | 6 (2, 11) | 8 (4, 15) | <0.001 | 8 (3, 16) | 11 (6, 20) | <0.001 |
|  | n=1032 | n=1093 |  | N=1110 | n=1204 |  |
| ESR, mL/hour | 48 (30, 71) | 47 (28, 77) | 0.8 | 59 (36, 93) | 62 (32, 95) | 0.9 |
|  | n=282 | n=297 |  | n=377 | n=424 |  |
| Neutrophil:lymphocyte ratio | 4.1 (2.5, 7.0) | 4.9 (3.0, 8.4) | <0.001 | 3.9 (2.5, 6.2) | 4.7 (3.0, 7.3) | <0.001 |
|  | n=1,238 | n=1330 |  | n=1255 | n=1339 |  |
| D-dimer, mg/L FEU | 0.91 (0.51, 2.0) | 0.88 (0.50, 1.9) | 0.8 | 1 (1, 3) | 1 (1, 4) | <0.001 |
|  | n=1042 | 1105 |  | n=1128 | 1209 |  |
| Ferritin, ng/mL | 348 (156, 735) | 767 (413, 1,362) | <0.001 | 463 (200, 945) | 954 (510, 1,739) | <0.001 |
|  | n=989 | n=1046 |  | n=1083 | n=1169 |  |
| IL-6, pg/mL | 33 (15, 74) | 53 (22, 122) | <0.001 | 39 (17, 99) | 60 (23, 154) | <0.001 |
|  | n=468 | n=504 |  | n=570 | n=641 |  |

**Supplemental Figure 1.**
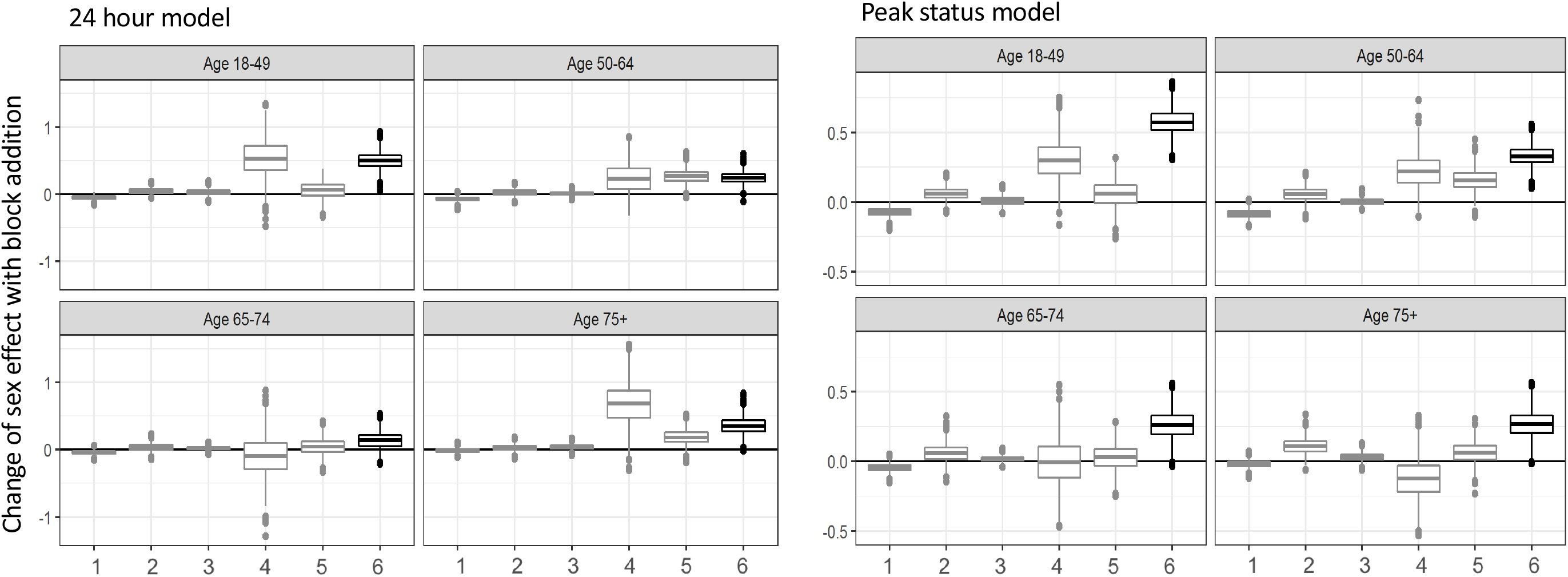
Change in log odds of the male sex effect associated with addition of variables blocks to the baseline model. Estimate of the difference in log odds with bootstrapped confidence intervals shown for each age group and variable block. The left panel is a model of severe/death outcome risk at 24hours, the right panel is from the models of peak status. Across all age groups, block addition modification of the baseline log odds was only significant for the inflammatory labs block (block 6) (24 hour model p<0.00001; peak status model p<1e10^−15^). Variable blocks as specified in the text: 1=BMI and admission source; 2=Comorbidities; 3=Smoking and alcohol use; 4=Presenting vitals; 5=General status labs; 6=Inflammatory labs.

## References

1. Jin J-M, Bai P, He W, et al. Gender Differences in Patients With COVID-19: Focus on Severity and Mortality. Frontiers in Public Health 2020; 8(152).

2. The Novel Coronavirus Pneumonia Emergency Response Epidemiology T. The Epidemiological Characteristics of an Outbreak of 2019 Novel Coronavirus Diseases (COVID-19) — China, 2020. China CDC Weekly 2020; 2(8): 113–22.

3. Salje H, Tran Kiem C, Lefrancq N, et al. Estimating the burden of SARS-CoV-2 in France. Science 2020; 369(6500): 208–11.

4. 5050 GH. https://globalhealth5050.org/covid19/ (accessed 05/02/2020.

5. Peckham H, de Gruijter NM, Raine C, et al. Male sex identified by global COVID-19 meta-analysis as a risk factor for death and ITU admission. Nat Commun 2020; 11(1): 6317.

6. Scully EP, Haverfield J, Ursin RL, Tannenbaum C, Klein SL. Considering how biological sex impacts immune responses and COVID-19 outcomes. Nat Rev Immunol 2020; 20(7): 442–7.

7. Bischof E, Wolfe J, Klein SL. Clinical trials for COVID-19 should include sex as a variable. J Clin Invest 2020; 130(7): 3350–2.

8. Mauvais-Jarvis F, Bairey Merz N, Barnes PJ, et al. Sex and gender: modifiers of health, disease, and medicine. Lancet 2020; 396(10250): 565–82.

9. Cornelius ME WT, Jamal A, Loretan CG, Neff LJ. Tobacco Product Use Among Adults — United States, 2019. MMWR Morb Mortal Wkly Rep 2020 2020; 69: 1736–42.

10. Key Substance Use and Mental Health Indicators in the United States:Results from the 2019 National Survey on Drug Use and Health. 09/2020 2020. https://www.samhsa.gov/data/report/2019-nsduh-annual-national-report (accessed 02/12/2021.

11. Wang QQ, Kaelber DC, Xu R, Volkow ND. COVID-19 risk and outcomes in patients with substance use disorders: analyses from electronic health records in the United States. Mol Psychiatry 2021; 26(1): 30–9.

12. Pun BT, Badenes R, Heras La Calle G, et al. Prevalence and risk factors for delirium in critically ill patients with COVID-19 (COVID-D): a multicentre cohort study. Lancet Respir Med 2021.

13. Klein SL, Flanagan KL. Sex differences in immune responses. Nat Rev Immunol 2016; 16(10): 626–38.

14. vom Steeg LG, Klein SL. SeXX Matters in Infectious Disease Pathogenesis. PLoS Pathog 2016; 12(2): e1005374.

15. Vom Steeg LG, Klein SL. Sex and sex steroids impact influenza pathogenesis across the life course. Semin Immunopathol 2019; 41(2): 189–94.

16. Scully EP. Sex Differences in HIV Infection. Curr HIV/AIDS Rep 2018; 15(2): 136–46.

17. Grebely J, Page K, Sacks-Davis R, et al. The effects of female sex, viral genotype, and IL28B genotype on spontaneous clearance of acute hepatitis C virus infection. Hepatology 2014; 59(1): 109–20.

18. Channappanavar R, Fett C, Mack M, Ten Eyck PP, Meyerholz DK, Perlman S. Sex-Based Differences in Susceptibility to Severe Acute Respiratory Syndrome Coronavirus Infection. J Immunol 2017; 198(10): 4046–53.

19. Karlberg J, Chong DS, Lai WY. Do men have a higher case fatality rate of severe acute respiratory syndrome than women do? Am J Epidemiol 2004; 159(3): 229–31.

20. Straub RH. The complex role of estrogens in inflammation. Endocr Rev 2007; 28(5): 521–74.

21. Schmiedel BJ, Singh D, Madrigal A, et al. Impact of Genetic Polymorphisms on Human Immune Cell Gene Expression. Cell 2018; 175(6): 1701–15 e16.

22. van der Made CI, Simons A, Schuurs-Hoeijmakers J, et al. Presence of Genetic Variants Among Young Men With Severe COVID-19. JAMA 2020.

23. Del Valle DM, Kim-Schulze S, Huang HH, et al. An inflammatory cytokine signature predicts COVID-19 severity and survival. Nat Med 2020; 26(10): 1636–43.

24. Takahashi T, Ellingson MK, Wong P, et al. Sex differences in immune responses that underlie COVID-19 disease outcomes. Nature 2020.

25. Klein SL, Pekosz A, Park HS, et al. Sex, age, and hospitalization drive antibody responses in a COVID-19 convalescent plasma donor population. J Clin Invest 2020.

26. Grzelak L, Velay A, Madec Y, et al. Sex differences in the decline of neutralizing antibodies to SARS-CoV-2. medRxiv 2020: 2020.11.12.20230466.

27. Chen Y, Klein SL, Garibaldi BT, et al. Aging in COVID-19: Vulnerability, immunity and intervention. Ageing Res Rev 2021; 65: 101205.

28. Richardson S, Hirsch JS, Narasimhan M, et al. Presenting Characteristics, Comorbidities, and Outcomes Among 5700 Patients Hospitalized With COVID-19 in the New York City Area. JAMA 2020.

29. Garibaldi BT, Fiksel J, Muschelli J, et al. Patient Trajectories Among Persons Hospitalized for COVID-19 : A Cohort Study. Ann Intern Med 2021; 174(1): 33–41.

30. PMAP: The Johns Hopkins Precision Medicine Analytics Platform. 2020. https://pm.jh.edu/. (accessed 02/12/21.

31. WHO. WHO R&D Blueprint novel Coronavirus COVID-19 Therapeutic Trial Synopsis.x02/18/2020 2020. https://www.who.int/publications/m/item/a-coordinated-global-research-roadmap (accessed 02/12/2021.

32. van Buuren S, Groothuis-Oudshoorn K. mice: Multivariate imputation by chained equations in R. Journal of Statistical Software 2011; 45(3): 1–67.

33. Computing RFfS. R Core Team (2021). R: A language and environment for statistical computing 2021. https://www.R-project.org/2021).

34. Peckham H, Gruijter N, Raine C, et al. Sex-Bias in COVID-19: A Meta-Analysis and Review of Sex Differences in Disease and Immunity. SSRN Electronic Journal 2020.

35. Vahidy FS, Pan AP, Ahnstedt H, et al. Sex differences in susceptibility, severity, and outcomes of coronavirus disease 2019: Cross-sectional analysis from a diverse US metropolitan area. PLoS One 2021; 16(1): e0245556.

36. Williamson EJ, Walker AJ, Bhaskaran K, et al. Factors associated with COVID-19-related death using OpenSAFELY. Nature 2020; 584(7821): 430–6.

37. Yoshida Y, Gillet SA, Brown MI, et al. Clinical characteristics and outcomes in women and men hospitalized for coronavirus disease 2019 in New Orleans. Biol Sex Differ 2021; 12(1): 20.

38. Petrilli CM, Jones SA, Yang J, et al. Factors associated with hospital admission and critical illness among 5279 people with coronavirus disease 2019 in New York City: prospective cohort study. BMJ 2020; 369: m1966.

39. Bi Q, Wu Y, Mei S, et al. Epidemiology and transmission of COVID-19 in 391 cases and 1286 of their close contacts in Shenzhen, China: a retrospective cohort study. Lancet Infect Dis 2020.

40. Kalish H, Klumpp-Thomas C, Hunsberger S, et al. Mapping a Pandemic: SARS-CoV-2 Seropositivity in the United States. medRxiv 2021.

41. Vashisht R, Patel A, Crews BO, et al. Age-and Sex-Associated Variations in the Sensitivity of Serological Tests Among Individuals Infected With SARS-CoV-2. JAMA Netw Open 2021; 4(2): e210337.

42. Bureau USC. https://www.census.gov/data/tables/time-series/demo/industry-occupation/median-earnings.html.

43. Boniol M, McIsaac M, Xu L, Wuliji T, Diallo K, Campbell J. Gender equity in the health workforce: analysis of 104 countries. Working paper 1. Geneva: World Health Organization; 2019.

44. Thompson AE, Anisimowicz Y, Miedema B, Hogg W, Wodchis WP, Aubrey-Bassler K. The influence of gender and other patient characteristics on health care-seeking behaviour: a QUALICOPC study. BMC Fam Pract 2016; 17: 38.

45. Barber SJ, Kim H. COVID-19 Worries and Behavior Changes in Older and Younger Men and Women. J Gerontol B Psychol Sci Soc Sci 2020.

46. Alkhouli M, Nanjundappa A, Annie F, Bates MC, Bhatt DL. Sex Differences in Case Fatality Rate of COVID-19: Insights From a Multinational Registry. Mayo Clin Proc 2020; 95(8): 1613–20.

47. Group RC, Horby P, Lim WS, et al. Dexamethasone in Hospitalized Patients with Covid-19 - Preliminary Report. N Engl J Med 2020.

48. Horby PW, Pessoa-Amorim G, Peto L, et al. Tocilizumab in patients admitted to hospital with COVID-19 (RECOVERY): preliminary results of a randomised, controlled, open-label, platform trial. medRxiv 2021: 2021.02.11.21249258.

49. Meier A, Chang JJ, Chan ES, et al. Sex differences in the Toll-like receptor-mediated response of plasmacytoid dendritic cells to HIV-1. Nat Med 2009; 15(8): 955–9.

50. Souyris M, Cenac C, Azar P, et al. TLR7 escapes X chromosome inactivation in immune cells. Sci Immunol 2018; 3(19).

51. Ursin RLK, S. L. Sex differences in respiratory viral pathogenesis and treatments. Annual Review of Virology 2021; in press.

